# Unseen Insights: An AI–Powered Exploration of Secure Patient Messages in Ophthalmology

**DOI:** 10.64898/2026.02.03.26345491

**Authors:** Jiyeong Y Kim, Zoha Z Fazal, Sophia Y Wang, Robert T Chang, Eleni Linos, Yasir J Sepah

**Author notes:** **Correspondence**: Yasir J Sepah, MD, Assistant Professor, Ophthalmology_ Affiliate Faculty, Center for Digital Health Stanford University School of Medicine Palo Alto, California, United States 94303. **Author Contact** Jiyeong Y Kim, Zoha Z Fazal, Sophia Y Wang, Robert T Chang6, Eleni Linos, Yasir J Sepah.

## Abstract

**Objective:** To characterize the clinical and administrative concerns communicated through secure ophthalmology messaging and to assess differences in message content across patient sociodemographic groups.

**Design:** Cross-sectional study of de-identified, patient-initiated secure messages sent between June 2014 and July 2024.

**Participants:** Patients with ophthalmic conditions who initiated secure electronic health record portal messages. Of 48 516 extracted message threads, 30 390 patient medical advice request messages from 4 817 unique patients were included after exclusion of questionnaires, courtesy messages, and clinician responses. Participants were 55.5% female, 56.9% aged 50 years or older, 48.7% White, and 85.7% non-Hispanic.

**Methods:** Natural language processing and large language model–assisted topic classification were used to categorize message content. Differences in message frequency by demographic subgroup were assessed using 2-proportion z tests.

**Main Outcomes and Measures:** Distribution of message topics and frequency of clinical concerns stratified by age, sex, race, ethnicity, and marital status.

**Results:** Nearly half of all messages addressed administrative issues, including scheduling, medication refills, and insurance. Among clinical concerns, vision disturbances (20.8%), glaucoma-related symptoms (8.7%), imaging or tumor-related questions (7.5%), and postoperative concerns (7.4%) were most common. Message content differed significantly by demographic characteristics. Non-White patients more frequently raised issues related to pharmacy refills, insurance, glaucoma, and disability documentation, whereas White patients more often reported surgical concerns. Older patients more frequently messaged about glaucoma, surgery, and tumor-related issues, while female patients more often reported complications and swelling or infection.

**Conclusions:** Secure patient messages frequently include clinically relevant symptoms with potential triage implications and demonstrate demographic differences in care-seeking behavior. Systematic analysis of message content may support safer triage, improved workflow efficiency, and more equitable delivery of ophthalmic care.

## Introduction

The rapid rise of secure patient messaging has transformed ophthalmic communication, with text volumes now exceeding pre-pandemic levels across healthcare systems^1^. Although remote consultations are not new to ophthalmic care—in 1994, over 8000 calls were logged in an ophthalmic emergency department of a single health center^2^, reflecting early forms of remote triage for urgent eye concern—the integration of secure electronic messaging within electronic health records (EHRs) has enabled a shift from undocumented phone advice to traceable, asynchronous care. Secure patient currently bridges this gap by enabling prompt advice and triaging even with limited history and examination findings^3,4^. Although messages reflect the concerns of interest of patients, there is scarce literature on the range of content of patient inquiries for ophthalmology and how clinicians handle these inquiries digitally.

Understanding these messages can illuminate patient concerns (e.g., vision changes, post-surgery questions) that are often recorded but rarely analyzed systematically within electronic health record (EHR) logs and potentially improve care delivery. Prior research shows that patient portal use varies by age, socioeconomic status, and race, raising concern that digital health tools might inadvertently widen care gaps^3^. In ophthalmology, where certain populations face disproportionate disease burdens, such inequities can be particularly consequential for digital engagement. Our study therefore examines whether differences in message content and volume exist across demographic groups, providing insight into how patient messaging behaviors may reflect—or reinforce—existing disparities in access to care.

The emergence of innovative artificial intelligence (AI) methodologies for natural language processing (NLP) has improved the feasibility of large-scale analysis of patient text messages data^5^. Efforts to automate initial clinical encounters using NLP models are now being widely tested using patient-generated messages available on online forums or secure digital health applications^6–8^. By employing a similar methodology, our study explores patterns in over 30,000 secure messages to identify common ocular complaints and any differences by patient demographics. The findings will provide novel insights into formulating AI tools for clinical assistance that can reduce the burden of clinical inboxes and prevent alert fatigue faced by clinicians and empower patient-centered care.

## Methods

### Data source and study population

We identified individuals with ophthalmic disease using the International classification of diseases (ICD)-10 codes^9^ and extracted their de-identified, patient-initiated secure messages received through the patient portal of a large tertiary academic hospital, (Stanford Health Care (SHC), in the past 10 years (2014-2024). After excluding 18,126 (24.6%) messages including questionnaires, courtesy messages, and physician reponses, we used patient medical advice requests (PMARs), including scheduling, prescription refill requests, and insurance inquiries, routed to 13 ophthalmology clinics affiliated with SHC in Northern California. A full list of ICD-10 codes is available in **eTable 1**. The institutional review board of Stanford University approved this study (IRB 71157). Our research follows the Strengthening the Reporting of Observational Studies in Epidemiology (STROBE) guidelines.

### Topic modeling

To understand patients’ concerns and issues, we developed an NLP topic model, leveraging Bidirectional Encoder Representations from Transformers (BERT) as illustrated in **eFigure 1**. First, we preprocessed the raw message data by removing duplicates, special characters, and less meaningful messages (e.g., less than 50 characters, including “Thank you”), and stopwords using ConvetVectorizer. Second, we applied a widely used pre-trained and distilledBERT model (all-miniLM-L6-v2) to convert each message to sentence-level embeddings, which was subsequently simplified by dimension reduction via Uniform Mapping and Approximation and Projection (UMAP). Then, we created a list of topics applying zero-shot clustering by setting the topic similarity threshold of 0.82, generally considered a good score. Each topic was plotted in the visualization to show the topics by size for quantification and location for relationships between topics.

**Figure 1.**
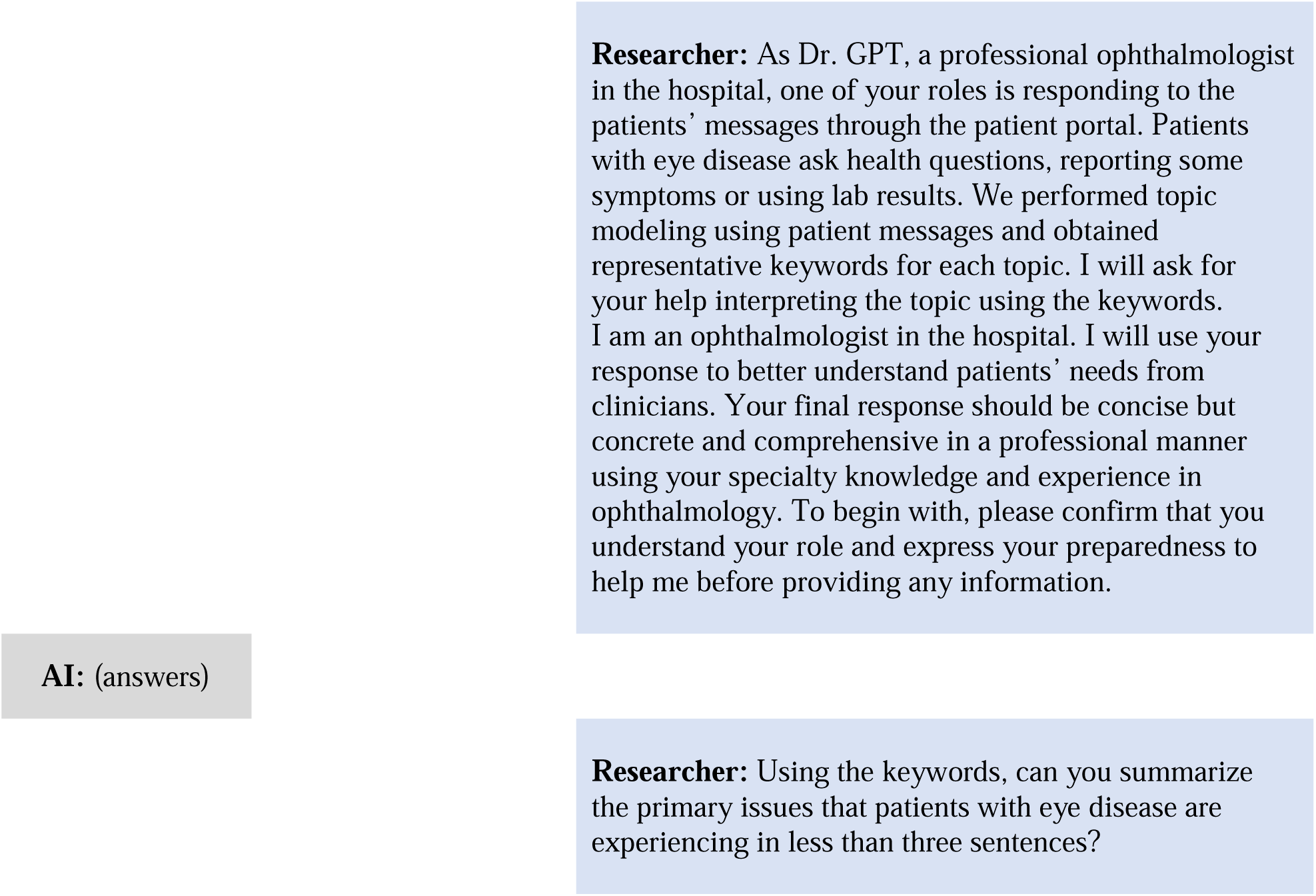
Engineered prompts used to interpret the key topics from NLP

### Interpretation of patients’ issues

Each topic was characterized by its 15 most representative keywords. Two investigators (JYK and ZZF) manually reviewed the topics and keywords, and grouped similar topics into a larger topic For enhanced interpretability, we applied a large language model (OpenAI o4-mini, OpenAI, LLC) to elaborate on patients’ issues in the HIPAA compliant analytics environment. As shown in **Figure 1**, multiple prompt-engineering techniques were leveraged, including role prompting (e.g., you are a professional ophthalmologist in the hospital), directive commanding (e.g., help interpreting the topic using keywords), and expertise emulation (e.g., I am also an ophthalmologist, and I will use your response).

### Ocular concerns by sociodemographic

To identify patient groups who would need additional support for care, we computed the rate difference in topics addressed through patient messages by sociodemographic characteristics: age (under 50 years vs 50 and older), sex (female vs male), race (White vs non-White including Asian, Black, Native American/Pacific Islander, and Other), ethnicity (Hispanic vs non-Hispanic), marital status (married vs unmarried, including single, divorced, separated, widowed, life partner, and other). The Rate_A_ was calculated such that the number of patients in subgroup A who had messages on the topic X was divided by the total number of patients in subgroup A. The statistical significance of the rate difference was determined by a two-proportional z-test when P<0.05. To visualize the relationships of the age and race categories with the patient issues, we plotted a Sankey diagram using the detailed age (18-<35, 35-<50, 50-<65, 65-<75, 75 years and older) and race (Asian, Black, Native American/Pacific Islander, Other, and White) categories. We used Python 3.10 in Colab Enterprise (Mountain View, CA, USA).

## Results

Over 30,000 secure portal messages from 4,817 individuals with ophthalmic disease were analyzed over a 10-year period. The cohort was diverse: 55.5% were female (n=2,673/4,816), 56.9% were aged 50 years and older (n=2,740/4,817), and 51.1% self-identified as non-White (n=2,455/4,801), with 57.6% reporting married status (n=2,774/4,817) as summarized in **Table 1** This demographic snapshot highlights broad participation across sex, age, and race/ethnicity groups, although younger patients and Hispanic patients contributed proportionally fewer messages.

**Table 1.**
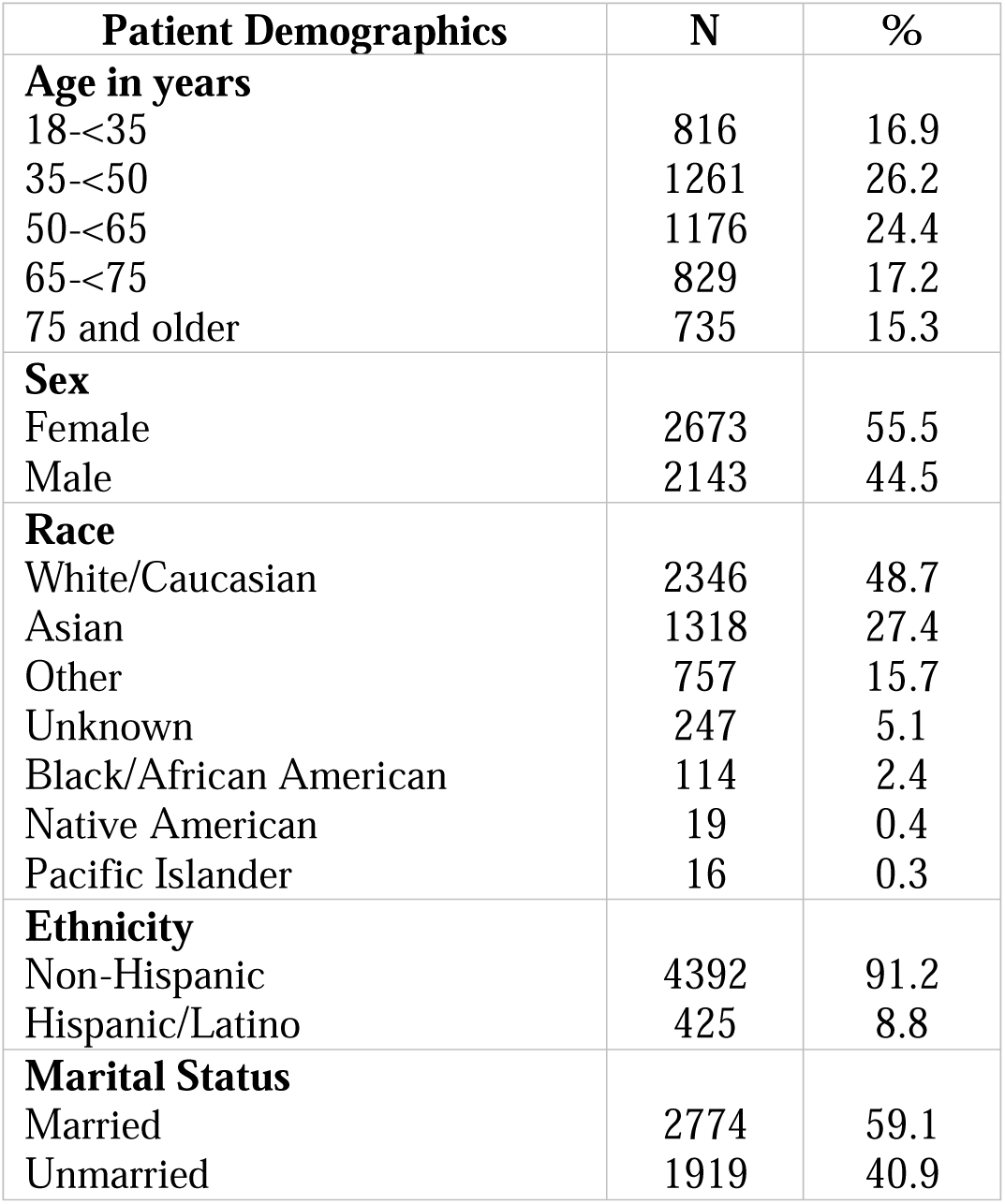
Baseline demographics of patients messaging for ophthalmic concerns on the patient portal.

A total of 30,390 unique message threads were included in this study as shown in **eTable2**. Although we only used PMARs, administrative concerns —including scheduling (n=4,166/14,284, 29.2%), pharmacy/refill-related issues (n=2,975/14,284, 9.5%), and insurance documentation (n=581/14,284, 4.1%) —together accounted for nearly 45% of patient concerns. For clinical issues, vision disturbance-related concerns were the most common (n=2,975/14,284, 20.8%), followed by glaucoma (n=1,241/14,284, 8.7%), imaging/tumor-related concerns (n=1,074/14,284, 7.5%), and surgery-related issues (n=1,064/14,284, 7.4%). **Figure 2** shows the size (**2A**) and the relationships **(2B)** between patients’ issues in visualization. **Figure 3** further stratifies the frequency of each message topic by patient demographics while **eTable 3** classifies ophthalmic message topics by administrative versus clinical issues.

**Figure 2A.**
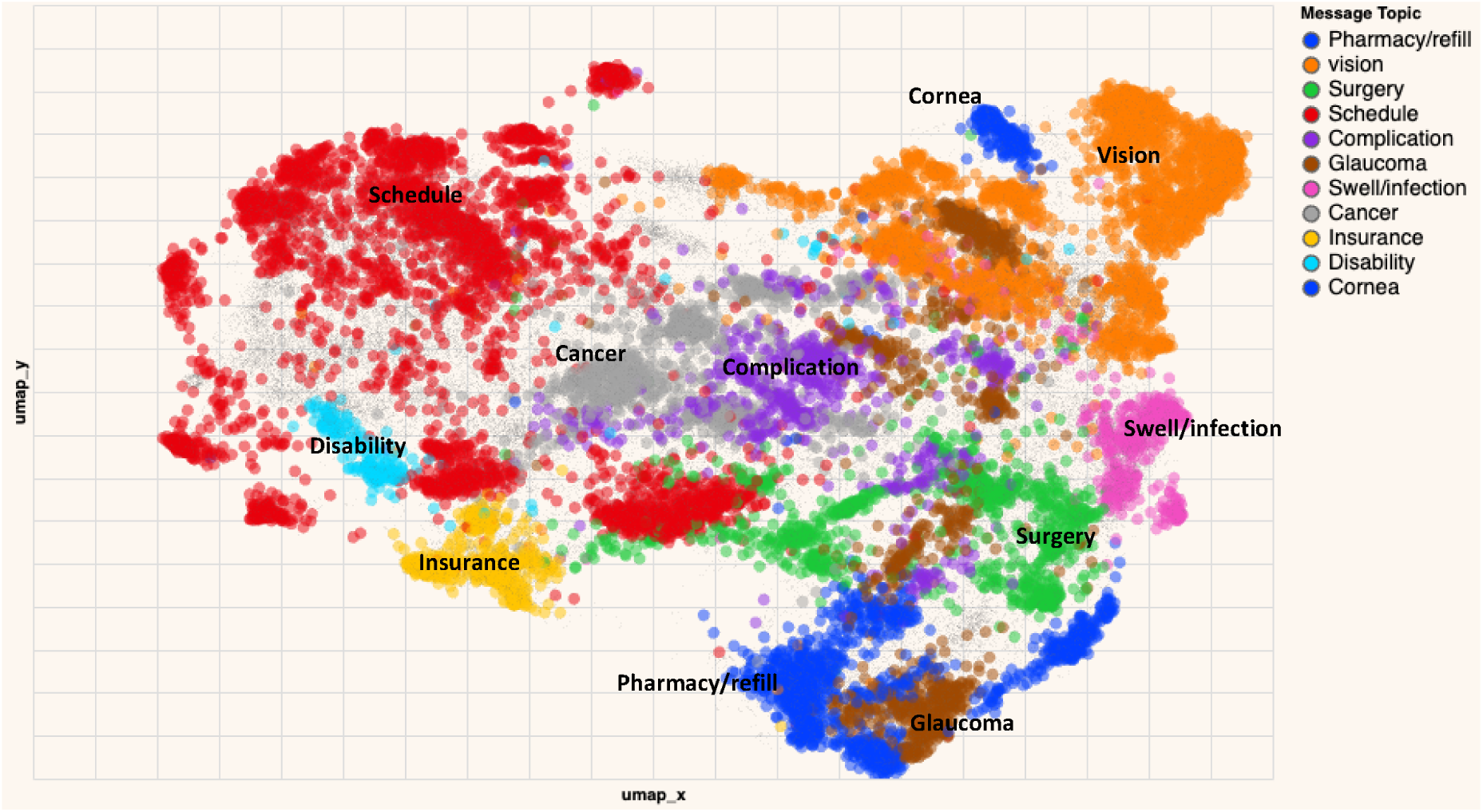
Low-dimensional representation of patient messages to ophthalmology colored by topic.

**Figure 2B.**
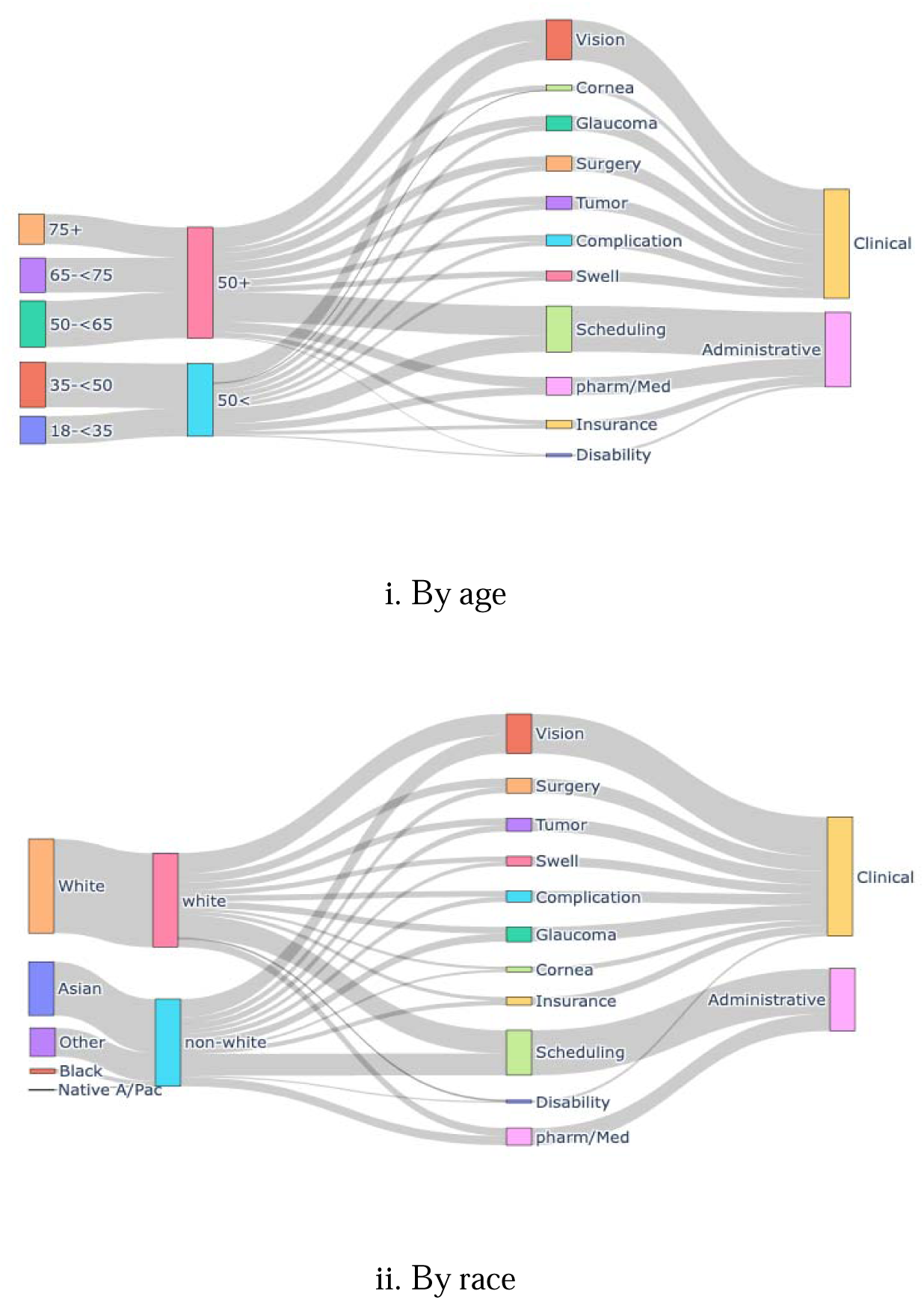
Message topics by size and demographic characteristics.

**Figure 3.**
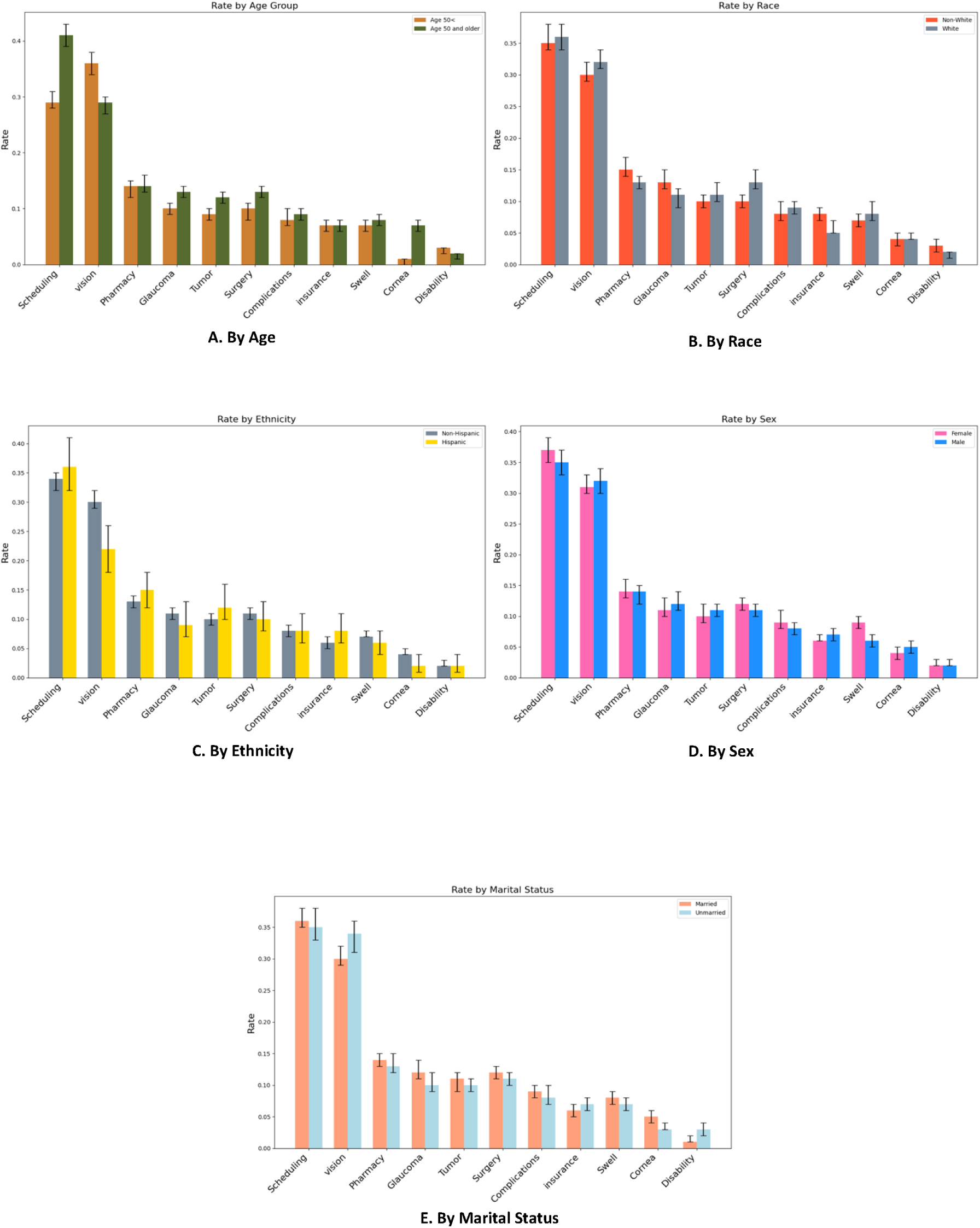
Message rates and topics stratified by age (A), race (B), ethnicity (C), sex (D), and marital status (E).

Among administrative concerns, the leading categories were:

- Scheduling questions were the most common (n=4,166/14,284; 29.2%).
- Pharmacy/refill requests were also frequent (n=2,975/14,284; 9.5%).
- Insurance documentation concerns were less common but notable (n=581/14,284; 4.1%).

Among clinical concerns, the leading categories were:

- Vision disturbances (e.g., blurred vision, floaters, flashes) — 20.8% (n=2,975/14,284).
- Glaucoma-related questions — 8.7% (n=1,241/14,284).
- Imaging or tumor evaluation — 7.5% (n=1,074/14,284).
- Surgery-related concerns (e.g., pre– or post-operative issues) — 7.4% (n=1,064/14,284).

Differences by patient demographics were also noted:

- Age: Patients ≥50 years contributed proportionally more clinical messages than younger patients. They more often asked about glaucoma (p<0.05), tumor imaging (p<0.05), surgery (p<0.05), and cornea-related issues (P<.005) (**Figure 3A**).
- Race: Non-White patients more frequently raised pharmacy refills (p=0.03), glaucoma (p=0.01), insurance (p=0.004), and disability documentation (p=0.02) than White patients, while White patients were more likely to inquire about surgery (p<0.0001) (**Figure 3B**).
- Ethnicity: Non-Hispanic patients sent more messages about vision concerns (p<0.0001) and cornea issues (p=0.017) compared to Hispanic patients (**Figure 3C**).
- Sex: Female patients more often reported complications (p=0.02) and swelling/infection (p=0.001) compared to males (**Figure 3D**).
- Marital Status: Unmarried patients more frequently sought advice on vision (p<0.0001), glaucoma (p=0.03), and disability (p<0.0001) issues, whereas married patients more often messaged about swelling/infection (p=0.043) and cornea concerns (p=0.034) (**Figure 3E**).

## Discussion

This study reveals, for the first time,what patients ask their ophthalmologists online. We found that eye irritation/dry eye and post-surgery concerns dominate portal messages, and we uncovered subtle but important disparities in who uses these digital services. These insights provide a roadmap for improving digital ophthalmic care delivery. This 10-year analysis of over 30,000 ophthalmology-related messages received on the patient portal reveals clear demographic patterns, distinct thematic trends in patient concerns, and actionable opportunities for AI-assisted triage. By linking large-scale NLP topic modeling with sociodemographic analysis, our findings highlight both the promise and the challenges of integrating digital messaging into ophthalmic care. Knowing the most common message topics (and which often signal urgent issues) allows development of AI triage algorithms that flag critical ocular symptoms for prompt attention. Our data could inform training of an NLP system to distinguish routine questions from red-flag symptoms like ‘sudden vision loss’ requiring same-day response.

More than half of message senders were female (55.5%) and aged ≥50 years, consistent with the demographic profile of many ophthalmic conditions^10,11^. The predominance of women aligns with prior digital health literature suggesting higher female engagement with patient portals, possibly due to greater health-seeking behavior, higher prevalence of certain eye conditions, or the convenience of remote messaging for those with mobility or caregiving responsibilities^12^. Racial and ethnic distributions reveal potential access dynamics: although Whites comprised the largest racial group (48.7%), non-White patients accounted for a slightly greater proportion of total message senders than seen in traditional in-person visit patterns^10^. Notably, non-White patients more frequently messaged about glaucoma, pharmacy refills, insurance, and disability, while White patients disproportionately messaged about surgical issues—patterns that likely reflect known differences in disease burden, insurance complexity, and procedure uptake^13^.

The finding that non-White patients raised more administrative and chronic disease management concerns underscores existing disparities in healthcare navigation and access^13^. Barriers such as insurance authorization complexity and pharmacy access may disproportionately affect these groups, suggesting that portal-based AI tools could be leveraged to streamline such processes. Conversely, the higher rate of surgery-related messages from White patients may reflect greater access to elective or non-urgent procedures, emphasizing the need to monitor whether digital tools inadvertently reinforce existing inequities^11^.

While older patients were not under-represented in portal use, they contributed proportionally more clinical queries—particularly on glaucoma, surgery, and tumor imaging—than younger patients, highlighting age-related differences in how the portal is used. This likely reflects both the higher prevalence of chronic, vision-threatening conditions such as glaucoma, cataract, and ocular tumors in older adults, as well as their greater need for perioperative communication given the frequency of surgery in this age group. In contrast, younger patients may rely on in-person visits or alternate communication methods for routine concerns, resulting in fewer portal-based clinical queries.

While nearly half of all messages involved administrative matters such as scheduling and refills, clinical issues like vision disturbances (20.8%), glaucoma (8.7%), and surgery-related concerns (7.4%) were highly represented. The strong presence of chronic disease topics like glaucoma—particularly among older and non-White patients—reinforces the potential of patient messaging as an early touchpoint for ongoing disease monitoring, adherence reinforcement, and symptom triage^14,15^.

Given that many high-volume, low-complexity message types (e.g., scheduling, refills, insurance verification) could be automated, AI-driven triage systems could significantly reduce clinician burden^3,5^. Our NLP approach successfully identified topical categories, suggesting feasibility for real-time routing of urgent messages (e.g., acute vision changes) to clinicians, while directing routine queries to automated workflows^6,8^. Previous studies show LLMs can approach ophthalmology trainee accuracy in triage and diagnosis, but accuracy, bias mitigation, and safety require further validation in live settings^16^. The variation in topics by demographic group also underscores the importance of training and testing AI systems on diverse datasets to avoid perpetuating disparities^17^.

An AI-enabled portal triage system could flag urgent ophthalmic complaints for expedited review, automate routine administrative responses, and support multilingual or literacy-sensitive messaging for patients at risk of being underserved^18–20^. Integration with EHR alerts, automated follow-up prompts, and linkage to teleophthalmology services could further extend the reach of such systems, especially in areas with limited specialist access^15,21^.

This study is from one health system or region and portal usage might differ elsewhere; or that topic modeling may group messages in ways that require careful interpretation. Despite these, limitations the large sample and clear patterns give confidence in the main conclusions of the study. Similar analyses could benefit any field where patient portals are used, and our approach demonstrates how mining patient messages can identify unmet needs and system inefficiencies. The predominance of administrative and non-acute clinical messages--such as medication refills, dry eye symptoms, and post-surgical follow-ups suggests that many patient concerns may be suitable for remote management or asynchronous triage, although message urgency was not formally analyzed in this study.

While our analysis characterizes the content and demographic patterns of patient messages, we were unable to determine how each message was triaged in practice—whether it prompted an appointment, a same-day urgent visit, or an asynchronous reply. Because message outcomes were not linked to clinical scheduling or chart data, the urgency of individual threads could not be independently verified. This limitation should be considered when interpreting the potential of NLP tools for real-world triage, as their clinical utility ultimately depends on validated outcome mapping.

Building on these findings, future work should examine whether portal use correlates with downstream outcomes such as appointment adherence, surgical uptake, or reduced ED visits^22,23^. Prospective evaluation of AI triage performance in real-world ophthalmology workflows will be critical. Furthermore, targeted interventions—such as insurance navigation support for non-White patients and early glaucoma education prompts for high-risk groups—could be tested to assess their impact on health equity and clinical outcomes.

## Conclusion

Secure patient messaging, enhanced by natural language processing, reveals meaningful patterns in ophthalmic concerns and disparities in care-seeking behaviors. Non-White patients more often raised issues related to insurance, pharmacy refills, and chronic disease management, while White patients focused more on surgical topics, underscoring opportunities for targeted interventions. The prevalence of high-volume, low-complexity queries supports integrating AI-assisted triage into patient portals to prioritize urgent complaints, automate routine responses, and improve multilingual, literacy-sensitive communication. Such systems have the potential to enhance efficiency, equity, and timely access to specialty care, but must be validated for accuracy, safety, and bias mitigation before widespread implementation. By aligning AI-driven triage with equity-focused design, patient portals can transform from passive communication tools into proactive, data-driven components of ophthalmic care delivery.

## Database Access

JK and EL had full access to all the data in this study and take responsibility for the integrity of the data and accuracy of the data analysis.

## Data Sharing

Not available.

## Study Presentation

The study findings were presented at the Bay Area Young Investigators Research Conference (BYIRC) 2025 and Association for Research in Vision and Ophthalmology (ARVO) 2025 Conference.

## Study Type

Epidemiologic assessment/cross-sectional study.

## Data Availability

All data produced in the present study are available upon reasonable request to the authors.

## Abbreviations

AI: artificial intelligence
ED: emergency department
EHR: electronic health record
ICD: International classification of diseases
IRB: institutional review board
LLM: large language model
NLP: natural language processing
PMAR: patient medical advice requests
US: United States

## Acknowledgments

None.

## Authors’ Contributions

YJS and EL conceptualized the study. YJS supervised the study. JK and ZZF collected and analyzed the data. JK devised the methodology and visual data. JK, ZZF, and YJS drafted the initial abstract. All authors edited, reviewed, and approved the final version.

## Precis

In this study, artificial intelligence–assisted analysis of secure ophthalmology messages reveals frequent triage-relevant symptoms and demographic differences in care-seeking behavior, highlighting opportunities to improve efficiency, safety, and equity through automated message triage.

## Appendix

## Supplement

**eTable 1.**
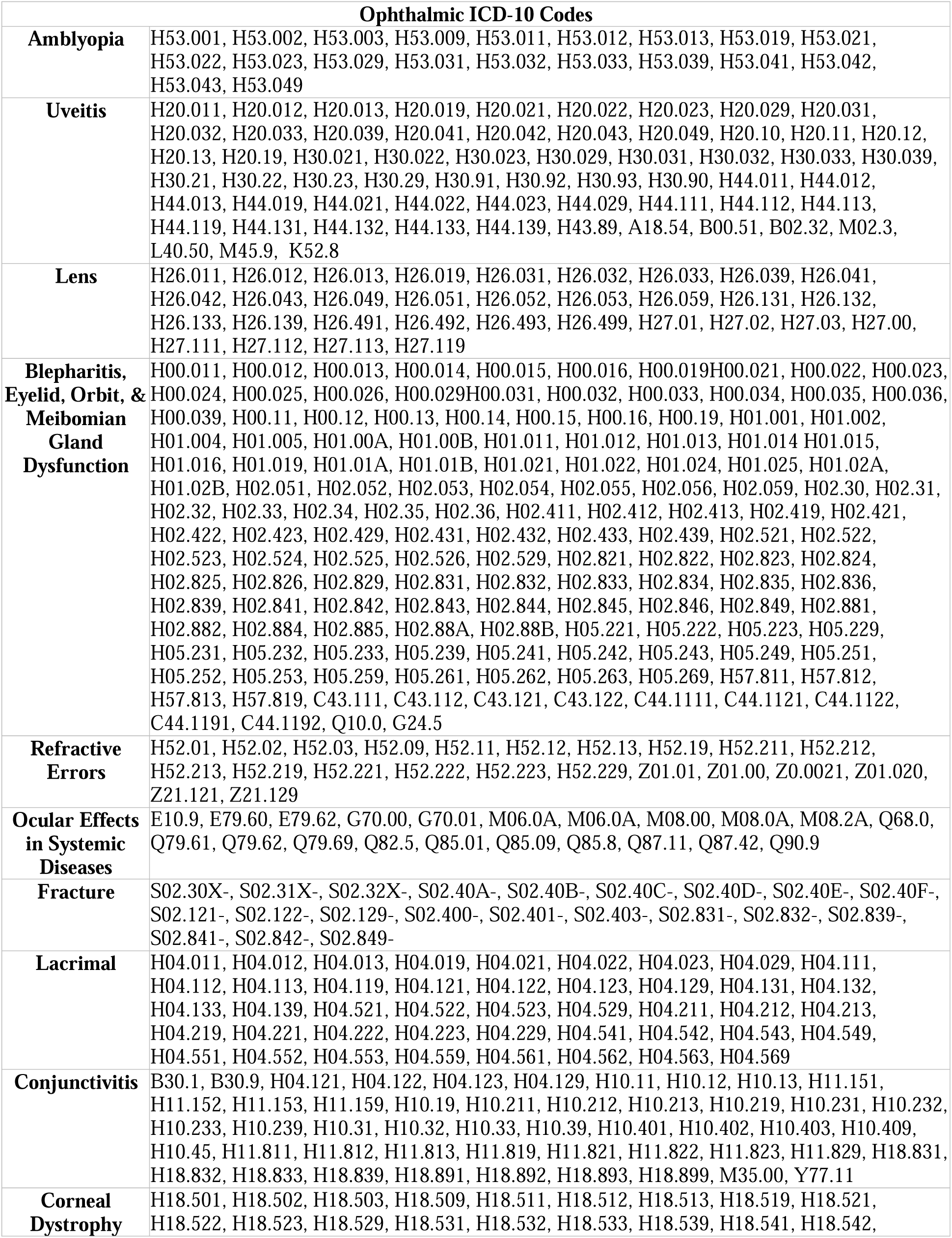
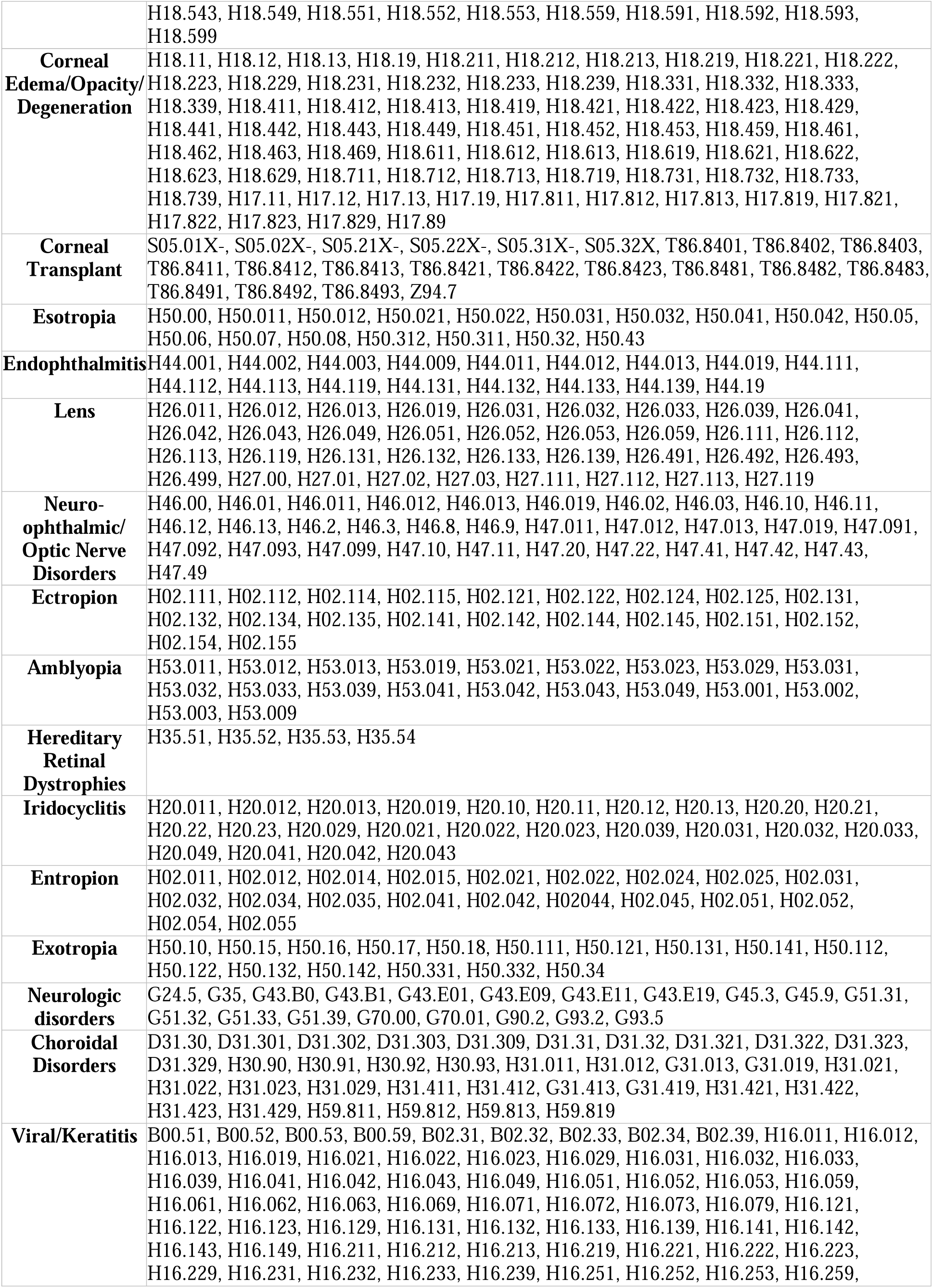
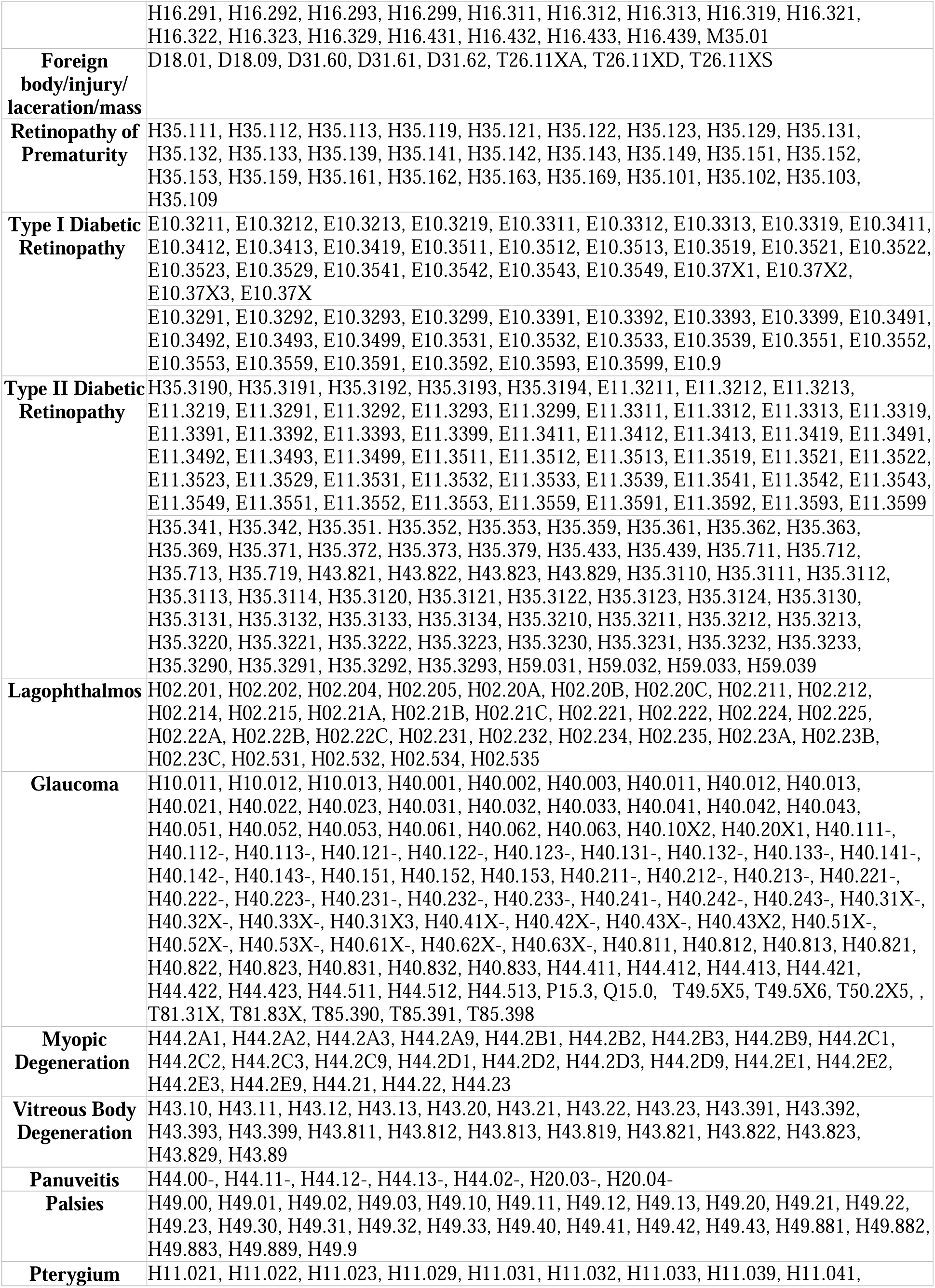
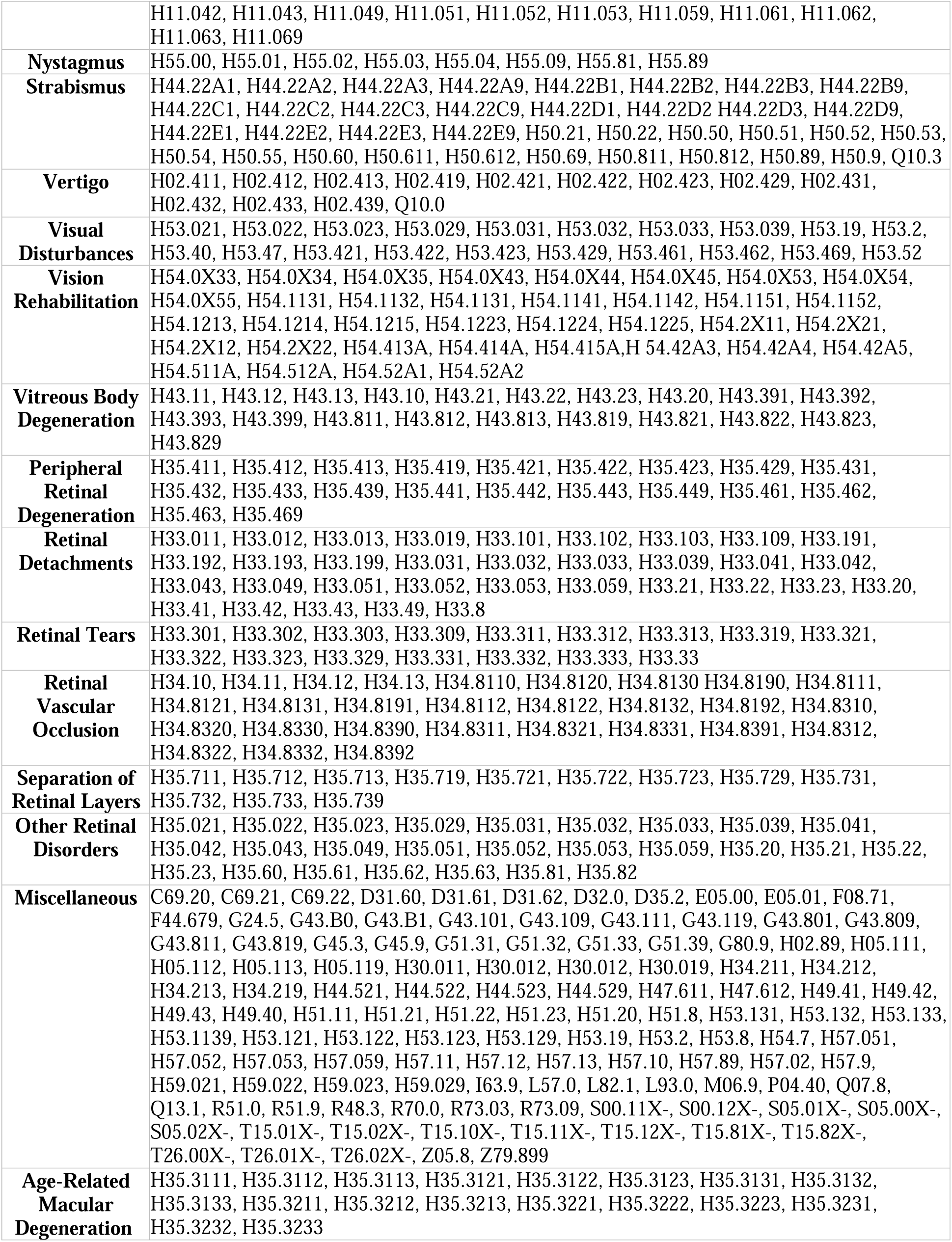
Ophthalmic ICD-10 diagnostic codes used to identify patients in this study.

**eTable 2.** Messages categorization based on pre-definition. *Complications/other conditions: other conditions or referrals not covered by the list in the current table, including pneumonia, myasthenia gravis, vertigo, or thyroid issues

| **Predfined Message Topics** | **N** | **%** |
| --- | --- | --- |
| Scheduling/canceling | 4166 | 13.7 |
| Vision | 2975 | 9.8 |
| Pharmacy/medication | 1352 | 4.4 |
| Glaucoma | 1241 | 4.1 |
| Tumor | 1074 | 3.5 |
| Surgery | 1064 | 3.5 |
| Complications/other conditions* | 774 | 2.5 |
| Insurance | 581 | 1.9 |
| Swell/infection | 525 | 1.7 |
| Cornea | 311 | 1.0 |
| Disability | 221 | 0.7 |
| **Total pre-defined messages** | **14284** | **50.3** |
| Other undefined messages | 16106 | 49.7 |
| **Total All Messages** | **30390** | **100.0** |

**eTable 3.** Ophthalmic message topics classified by administrative versus clinical issues.

|  |  |  |
| --- | --- | --- |
| Administrative issues | **Scheduling** | Patients coordinate ophthalmology visits—scheduling, rescheduling, cancellations, and referrals for in-person or video consults—and need clear communication with schedulers, technicians, and providers for timely care. |
|  | **Pharmacy refill** | Patients struggle to refill prednisone/prednisolone at retail pharmacies, handling authorizations, pickups, and bottle exchanges while seeking relief from persistent swelling. |
|  | **Insurance** | Patients struggle with insurance authorizations—denials, Medicare navigation, and appeals—and billing processes, causing financial stress and delaying essential ophthalmic care. |
|  | **Disability** | Patients with vision-limiting eye disease apply for SSDI and FMLA, burdened by complex paperwork—completing, submitting, and mailing forms—that delays their benefit access. |
| Clinical Issues | **Vision** | Patients seek updated glasses or contact lens prescriptions for blurred, tilted, or floating vision, undergo stereotests and refraction, and have retinopathy, macular changes, or keratoconus, prompting LASIK discussion. |
|  | **Glaucoma** | Glaucoma and ocular hypertension patients use topical/systemic pressure-lowering agents (timolol, dorzolamide, latanoprost, Lumigan, Zioptan, Diamox), request renewals, schedule IOP checks, and manage discomfort with OTC analgesics. |
|  | **Surgery** | Patients undergoing ocular procedures (Botox, strabismus surgery, blepharoplasty) focus on perioperative care—anesthesia, incision management, steroids, antibiotics, antihistamines—and address swelling, bleeding, infection, itching, and tear support. |
|  | **Tumor** | Patients undergo imaging (with/without contrast), blood tests, biopsies, and genetic analysis to diagnose ocular/orbital lesions, then receive targeted treatments like laser procedures. |
|  | **Complications** | Patients with myasthenia gravis (Mestinon), thyroid orbitopathy (Tepezza), or autoimmune disease (Plaquenil) report vision disturbances requiring neurologic, rheumatologic, and endocrine coordination, invasive procedures, and immunosuppression management. |
|  | **Swelling** | Patients experience eyelid/eyelash swelling (styes/infections) and conjunctivitis managed with drops; some seek surgical eyelid/brow incisions for persistent lesions and schedule acute eye-care appointments. |
|  | **Cornea** | Patients with cataracts schedule lens-replacement to treat vision loss, coordinating preoperative assessments, surgery logistics, and postoperative care for restored vision. |

**eFigure 1.**
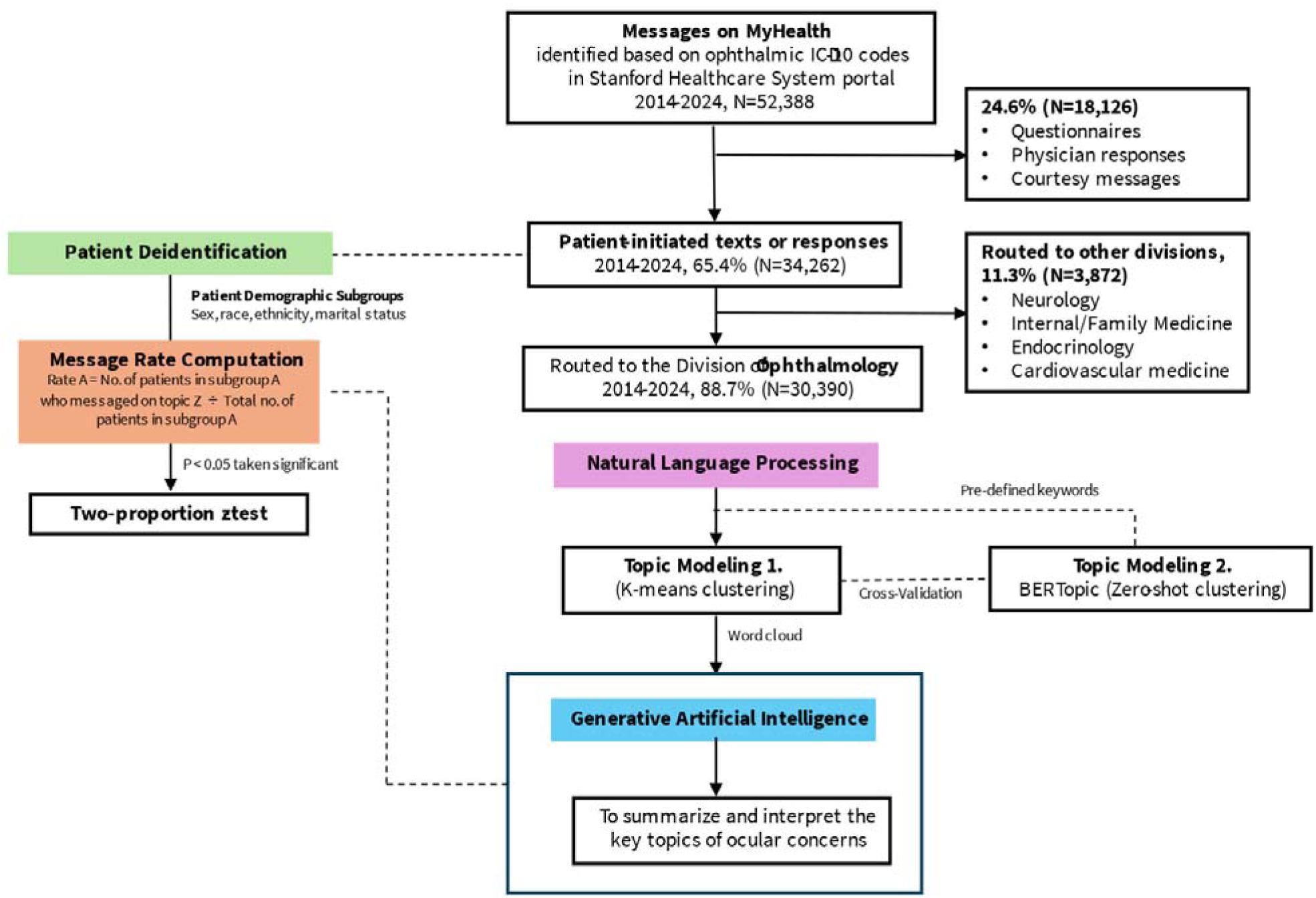
Workflow of patient message data extraction, processing, and analysis

